# Dermal trypanosomes in seropositive suspects of *gambiense* human African trypanosomiasis in Côte d’Ivoire

**DOI:** 10.1101/2025.04.08.25325162

**Authors:** Martial Kassi N’Djetchi, Mélika Barkissa Traoré, Innocent Abé, Bamoro Coulibaly, Valentin Nanan, Thomas Konan, Louis N’Dri, Ibrahim Sadissou, Jean-Mathieu Bart, Bruno Bucheton, Magali Tichit, David Hardy, Salimatou Boiro, Aïssata Camara, Christelle Travaillé, Aline Crouzols, Nathalie Petiot, Adeline Ségard, Lingue Kouakou, Mariame Camara, Mamadou Camara, Dramane Kaba, Mathurin Koffi, Vincent Jamonneau, Brice Rotureau

## Abstract

In the population at risk of *gambiense* human African trypanosomiasis (gHAT), the prevalence of extravascular parasite carriage remains unclear. Here, we conducted an observational clinical study in the hypo-endemic gHAT foci of Sinfra and Bonon in Côte d’Ivoire from 2019 to 2022. A total of 74 individuals were enrolled, including 45 suspects previously found positive at least once in a serological test for gHAT and followed by the national elimination programme of Côte d’Ivoire, as well as 29 seronegative controls. No significant differences between groups were observed for any epidemiological parameters and any clinical parameters at enrolment. Whereas trypanosome DNA was detected in the blood of 0/29 controls and 2/45 suspects, the presence of extravascular dermal trypanosomes was assessed by immuno-histochemistry (fixed trypanosome cells) and/or PCR (trypanosome DNA) in about 1/3 of the suspects (14/45, 31%). However, no *gambiense*-specific test was found positive in the present study. Hence, the skin could represent an anatomical reservoir for African trypanosomes sustaining a low level of transmission in hypo-endemic foci.

**Author summary:** In the population at risk of sleeping sickness, the number of people bearing trypanosome parasites in their skin remains unclear. Here, we conducted an observational clinical study in the hypo-endemic transmission foci of Sinfra and Bonon in Côte d’Ivoire from 2019 to 2022. A total of 74 individuals were enrolled, including 45 suspects previously found positive at least once in a test on blood and followed by the national elimination programme of Côte d’Ivoire, as well as 29 negative controls. No significant differences between groups were observed for any epidemiological parameters and any clinical parameters at enrolment. Whereas trypanosome DNA was detected in the blood of 0/29 controls and 2/45 suspects, the presence of dermal trypanosomes was assessed by immuno-histochemistry (fixed trypanosome cells) and/or molecular biology (PCR to detect trypanosome DNA) in about 1/3 of the suspects (14/45, 31%). However, no *gambiense*-specific test was found positive in the present study. Hence, the skin could represent an anatomical reservoir for African trypanosomes sustaining a low level of transmission in hypo-endemic foci.

## Introduction

With only 799 new cases reported in 2022, *gambiense* human African trypanosomiasis (gHAT or sleeping sickness), a neglected tropical disease caused by the tsetse-borne protist parasite *Trypanosoma brucei gambiense*, has been targeted for elimination (zero transmission) by 2030 by the World Health Organization (WHO) [1, 2]. Seven countries have already eliminated the disease as a public health problem, including Côte d’Ivoire in 2020, with nine gHAT cases detected in two health districts between 2015 and 2019 [3]. Then, between 2020 and 2022, only one gHAT case was detected in Côte d’Ivoire (Franco et al., 2024). These results offer encouragements towards the elimination of transmission by the end of 2025 on condition of maintaining a multidisciplinary one health approach and constant research activities to continuously adapt surveillance strategies in the epidemiological transition to zero incidence [3, 4]. In Côte d’Ivoire, as well as in any endemic areas, the recommended gHAT diagnosis algorithm relies on three successive steps (Table 1). First, a serological test on blood, such as the card agglutination test for trypanosomiasis (CATT) or a rapid diagnostic test (RDT), is performed [5]. Second, for seropositive subjects only, a parasitological confirmation is required to assess the presence of living trypanosomes in a biological fluid (blood or buffy coat or lymph node aspirate) under a microscope, usually after sample concentration with the mini-anion exchange centrifugation technique (mAECT) [6]. Third, if trypanosomes are detected in the blood, cerebrospinal fluid is sampled by lumbar puncture for searching for trypanosomes under a microscope to determine the disease stage and adapt the treatment accordingly. Seronegative individuals and seropositive subjects without parasitological confirmation are not treated.

**Table 1.**
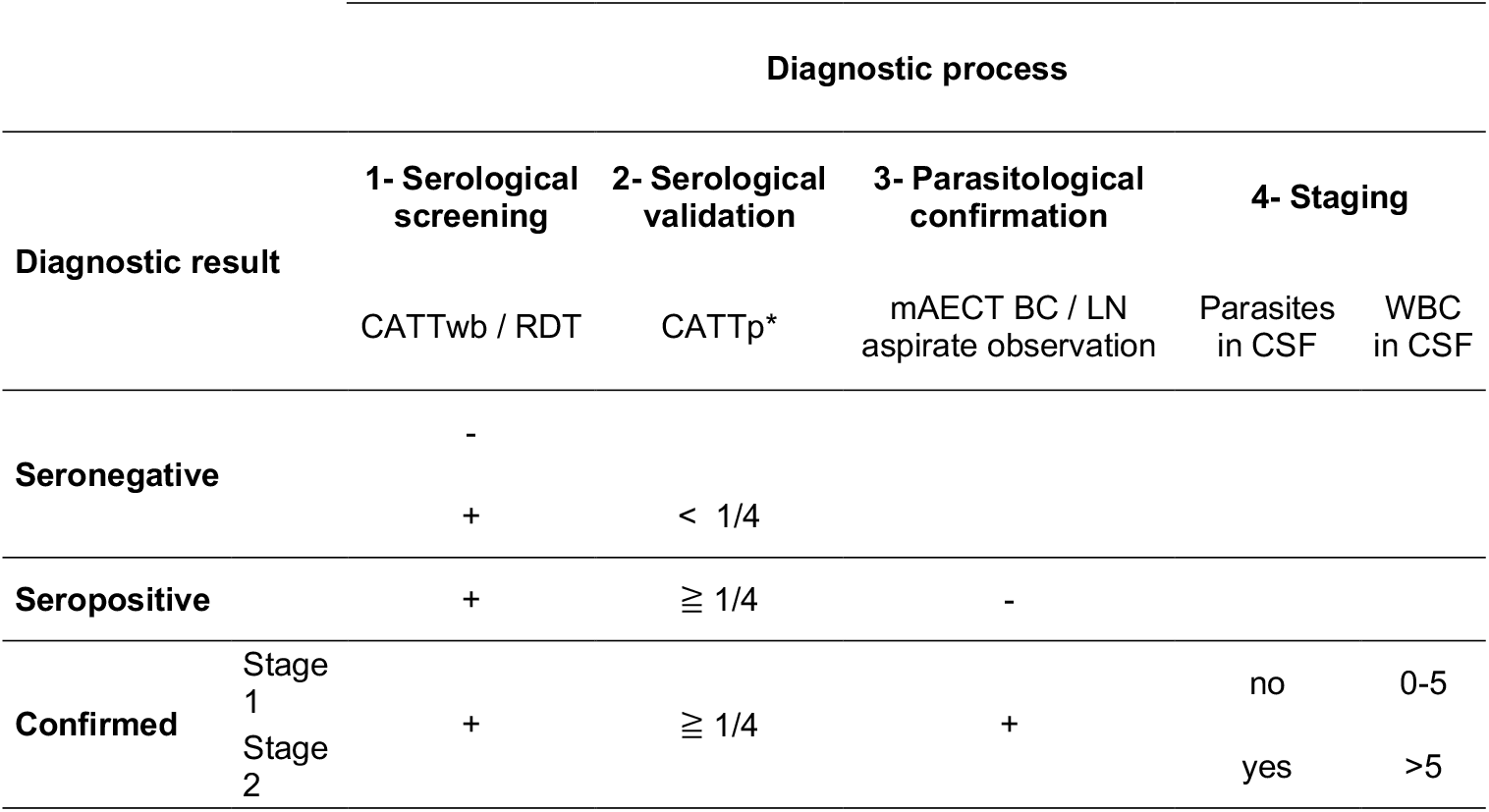
Diagnostic process. CATTwb / CATTp: card agglutination test for trypanosomiasis on whole blood / plasma; RDT: Rapid Diagnostic Test for HAT; mAECT BC / LN aspirate: mini anion-exchange column technique on buffy coat / lymph node aspirate; WBC: white blood cells; CSF: cerebrospinal fluid; *Highest plasma dilution with a positive result.

To move towards gHAT elimination, a combination of both exhaustive and targeted medical screening strategies based on this diagnostic algorithm, as well as vector control programs to reduce the risk of transmission in the most at-risk areas, were implemented in parallel [3]. Among the targeted medical strategies, a follow-up of unconfirmed seropositive subjects, considered as potential latent carriers, was also maintained to diagnose and treat all possible cases as well as to estimate the impact of persisting human reservoirs on the elimination goal [3, 7].

Indeed, the presence of *T. b. gambiense* parasites was reported in the extravascular compartment of the mammalian host skin, under experimental conditions in animal models [8, 9], and more importantly, during the natural progression of the disease in humans patients [10, 11]. Even in absence of detectable parasites in blood, substantial quantities of extravascular trypanosomes persist in the basal layer of the dermis and can be transmitted to the tsetse vector in experimental conditions [8]. The presence of long-term seropositive individuals in the two hypo-endemic foci of Sinfra and Bonon in Côte d’Ivoire prompted us to investigate the involvement of skin-dwelling trypanosomes in this phenomenon. To refine our understanding of the epidemiological importance of skin-dwelling trypanosomes in this context, we performed a prospective observational study in some of the seropositive subjects of the historical cohort of long-term seropositive individuals in Sinfra and Bonon [3, 7].

## Methods

### Ethical approval

All investigations were conducted in accordance with the Declaration of Helsinki and fulfil the STROBE criteria. Approval for this study was obtained from the Comité National d’Ethique des Sciences de la Vie et de la Santé de République de Côte d’Ivoire (authorization #111-19/MSHP/CNESVS-kp and amendments). Children under 16 years of age and pregnant women were excluded from the study. Each participant was informed about the study’s objectives in their own language and provided written informed consent. For participants between 16 to 18 years of age, informed consent was also obtained from their parents. The data of individual study participants were randomly anonymized with a 4-digit code at enrolment by the MD in charge of the medical campaign.

### Study enrolment, screening and group definition

All subjects enrolled in this study came from the hypo-endemic gHAT foci of the Sinfra and Bonon districts, which are located in the central west part of the Republic of Côte d’Ivoire (Figure 1) [3]. All subjects were enrolled in October 2019 during a medical survey performed by the HAT National Elimination Programme, according to WHO recommendations and as described previously (Table 1) [3, 12]. Then, a first follow-up visit was organized in April 2021 and a second one in June 2022 for seropositive subjects only (Table 2). Individuals were screened with the card agglutination test for trypanosomiasis using whole blood samples (CATTwb) at all time-points, and with the SD Bioline HAT 1.0 rapid diagnostic test (RDT) on blood at enrollment only (Table 1). For those individuals who tested positive in the CATTwb or RDT, 5 mL of blood were collected in heparinized tubes and a two-fold plasma dilution series was used to determine the CATT plasma (CATTp) end titer. All individuals with CATTp end titers of 1/4 or higher underwent a microscopic examination of lymph node aspirate, whenever cervical swollen lymph nodes were present. Blood samples of CATTp-positive individuals were also centrifuged to obtain the buffy coat layer, which was tested for trypanosomes using the mini-anion exchange centrifugation test (mAECT BC) [6]. If trypanosomes were detected using this test, the infected individual underwent a lumbar puncture and their disease stage was determined by searching for trypanosomes using the modified simple centrifugation technique for CSF and by white blood cell (WBC) counts [13]. *Gambiense* HAT patients were classified as being stage 1 (0-5 WBC/µl and absence of trypanosomes in CSF) or stage 2 (>5 WBC/µl and/or presence of trypanosomes in CSF) and were treated accordingly by the National Elimination Programme strategy (Table 1).

**Table 2.** Group definitions and number of subjects enrolled per visit and group. CATT: card agglutination test for trypanosomiasis; TL: immune trypanolysis test.

| Groups | Number of subjects |  |  | Status before 2019 |  |  |  | Clinical history |
| --- | --- | --- | --- | --- | --- | --- | --- | --- |
|  | Enrollement<br>2019 | Follow-up 1<br>2021 | Follow-up 2<br>2022 | CATT | TL | Parasitology | Treatment |  |
| **PCT** | 6 | 2 | 2 | + | + | + | + | Previous case confirmed and treated before 2019 |
| **PCU** | 7 | 5 | 4 | + | + | + | - | Previous case confirmed before 2019 but untreated |
| **SeroTL** | 19 | 9 | 5 | + | + | - | - | Previous seropositive in CATT and in TL at least once before 2019 but negative in parasitology |
| **SeroCATT** | 13 | 9 | 9 | + | - | - | - | Previous seropositive in CATT at least once before 2019 but negative in both TL and parasitology |
| **Control** | 29 | 0 | 0 | - | - | - | - | No gHAT history |
| **Total** | 74 | 25 | 20 |  |  |  |  |  |

**Figure 1.**
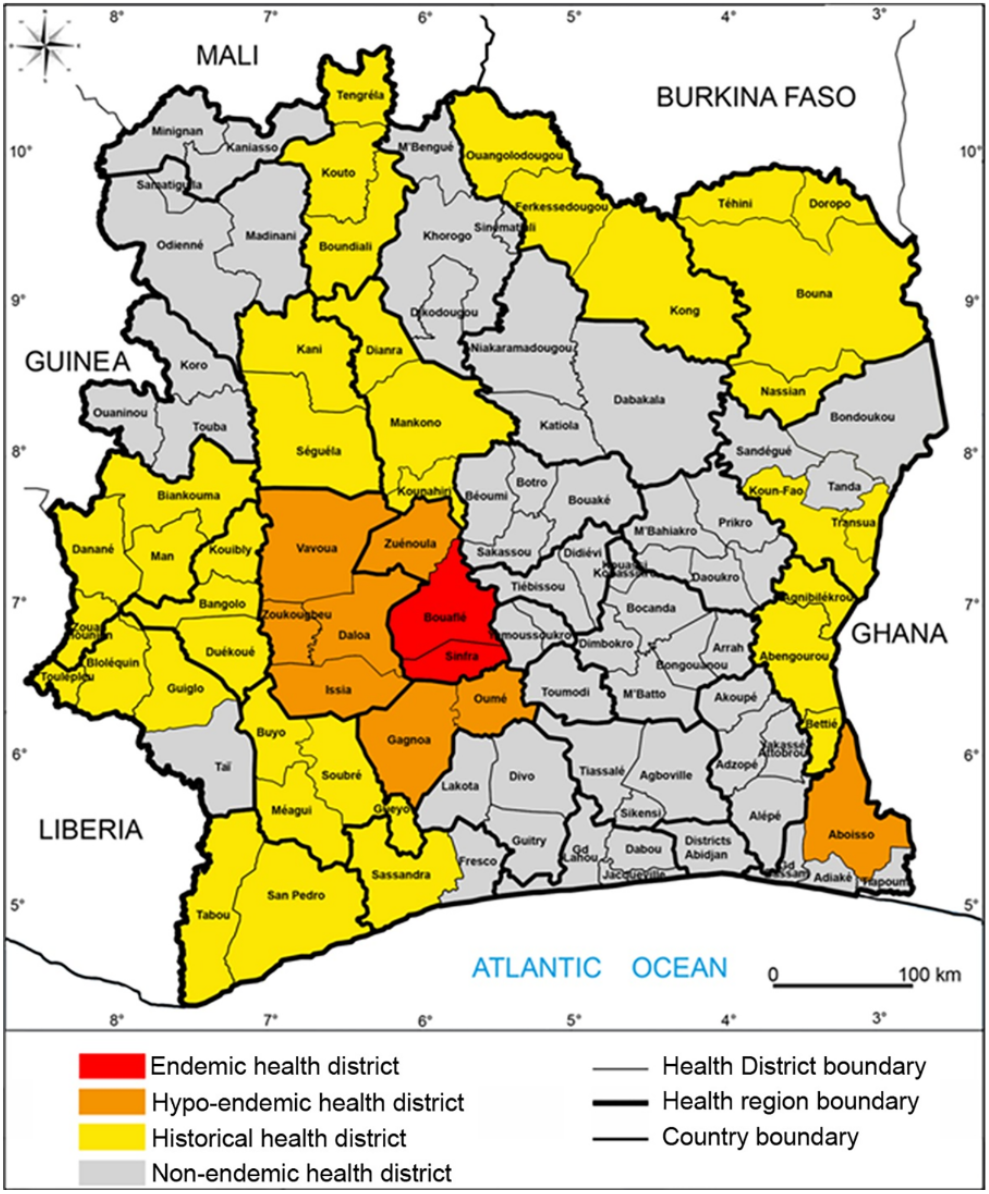
Map of the hypo-endemic gHAT foci in Côte d’Ivoire. This map shows the administrative districts of the Republic of Côte d’Ivoire. Hypo-endemic transmission foci in orange, old foci with last cases reported before 2019 in yellow (adapted from [3]).

Subjects were assigned to one of the 5 study groups according to their gHAT clinical history as defined in Table 2: previous cases confirmed and treated before 2019 (PCT), previous cases confirmed before 2019 but left untreated (refusal or lost-to-follow-up after diagnosis) (PCU), individuals found seropositive in CATT and in immune trypanolysis test (TL) at least once before 2019 but who remained negative in parasitology (SeroTL), and individuals found seropositive in CATT at least once before 2019 but negative in both TL and parasitology (SeroCATT). The clinical and diagnostic history of all seropositive individuals (PCT, PCU, SeroTL and SeroCATT) was available from the HAT National Elimination Programme records. In addition, seronegative controls with no gHAT history were randomly enrolled from the neighbourhood of the cohort subjects (Table 2).

### Field procedure and sampling

As in [10] and [11], at each visit, each participant underwent an epidemiological interview and a clinical examination, during which dermatological symptoms were assessed by a trained dermatologist. The following epidemiological data were collected: age (in years), sex, clinical history of HAT infections in the family since 2010, and occupational risk (occurrence of any regular activities such as farming, wood-cutting mining, hunting and fishing, during which an individual might be exposed to tsetse bites). The following general clinical data were also collected during the interview: fever, swollen cervical lymph nodes, weight loss, asthenia, eating disorders, sexual dysfunctions, repetitive headaches, circadian rhythm disruptions and/or any other behavioral changes during the last three months. Dermatological signs of pruritus (skin itch) and dermatitis (skin inflammation) were also investigated, and a careful examination of the entire body was performed to detect any symptoms that might be related to skin infections.

Finally, two superficial skin snip biopsies were sampled in sterile conditions from the right back shoulder of all enrolled subjects. In this study, the skin region was the same as in Camara *et al*. [10] for comparison, yet the choice of sampling superficial snip biopsies rather than deep punch biopsies was preferred by the HAT National Elimination Programme to limit sampling invasiveness and allow a more rapid healing of the lesion. Biopsies were performed under local anesthesia and were rapidly dressed. One first skin snip was rapidly fixed in 10% neutral buffered formalin at room temperature for immuno-histochemistry, and a second one was preserved in RNAlater at -20°C for molecular analyses. Plasma aliquots (0.5 mL) were also obtained from 10ml blood samples and preserved at -20°C in the field and -80°C in the lab for use in serological trypanolysis tests. An additional 0.5 mL blood sample was also collected preserved at -20°C for PCR.

### Immune trypanolysis test

A plasma sample from all participants was used to perform the immune trypanolysis test. This test detects complement-mediated immune responses activated by either the LiTat 1.3, LiTat 1.5 or LiTat 1.6 variable surface antigens specific for *T. b. gambiense*, as previously described [14, 15].

### Immunohistochemical detection of trypanosomes

Skin snip biopsy samples fixed in formalin and preserved at 4°C were trimmed and processed into paraffin blocks in the lab. Longitudinal sections of ∼2.5μm were prepared and processed using Dako Autostainer Plus (Dako, Denmark). Sections were immunolabelled with the *T. brucei*-specific anti-ISG65 antibody which targets the Invariant Surface Glycoprotein 65 (rabbit 1/800; gift from M. Carrington, University of Cambridge, UK) [16]. For immunolabelling, a horseradish peroxidase-coupled secondary antibody was used, and the staining was revealed with 3,3’- diaminobenzidine and counterstained with Gill’s hematoxylin. A non-infected West African skin specimen (Tissue Solutions Ltd., UK) and a *T. b. gambiense*-infected mouse skin specimen were also included with the samples as technical negative and positive controls, respectively. Immunostaining images were acquired using an automated Axio Observer Z1 microscope (Carl Zeiss, Germany) and analyzed using the ZEN 3.7 software (Carl Zeiss, Germany). The slides from each biopsy were blindly assessed by two readers. The positivity of a given skin-section slide was defined by the detection of at least five clearly morphologically distinguishable and antibody-stained trypanosomes in the most basal region of the dermis. In 2022, skin samples positive in IHC-ISG65 were confirmed in IHC-CRD with the *T. brucei*-specific anti-CRD antibody which targets the cross-reacting determinant (rabbit 1/750; gift from J. Bangs, University at Buffalo, USA) [17].

### TNA extraction

Skin snip biopsy samples in RNAlater and preserved at -20°C were treated for total nucleic acid (TNA) extraction in the lab. A non-infected West African skin specimen (Tissue Solutions Ltd., UK) and a *T. b. gambiense*-infected mouse skin specimen were also included with the samples as negative and positive technical controls, respectively. For blood samples, TNA extraction was performed on 0.5 mL blood aliquots. TNAs were extracted with DNeasy Blood and Tissue kits (Qiagen, Germany) following the manufacturer’s recommendations.

### Molecular detection of trypanosome TNA

For all blood and skin samples at all time-points, PCR-TBRN3 detection of *T. brucei s. l*. parasites was performed using primers targeting the DNA satellite repeat sequence (10,000 copies per cell) to generate a 168bp amplicon [5, 10]. For enrollment only, blood and skin samples were also tested in PCR-18S detecting *T. brucei s. l*. parasites [18], as well as in simple PCR-TgsGP directed against the single copy *TgsGP* gene [19].

### Data analyses

All data are available in Sup. Table 1. General descriptive analyses of anonymized data were performed using Excel 16.80 (Microsoft, USA). Statistical analyses were performed using Prism V10.1.1 (GraphPad, USA) software. For diagnostic parameters, differences between seronegative controls and seropositive individuals in each group were assessed using Fisher’s exact tests at 5% confidence.

## Results

### Population, follow-up and particular cases

A total of 74 subjects had no exclusion criteria and accepted to be enrolled in the study in 2019. Among them, 45 subjects belonging to the cohort followed up by the HAT National Elimination Programme (Table 2). These include 6 PCT, 7 PCU, 19 SeroTL and 13 SeroCATT. In addition, 29 seronegative controls with no gHAT history were randomly enrolled for the first visit only (Table 2). In 2021 and 2022, respectively 25/45 (56%) and 20/45 (44%) cohort subjects were followed up (Table 2).

As shown in Table 3, no significant differences between groups were observed for any epidemiological parameters and any clinical parameters at enrolment.

**Table 3.** Epidemiological and clinical characteristics of subjects. For each group (defined in Table 2) and each parameter, numbers represent the number of positive outcomes for a given parameter out of the total number of available results for this parameter, except for the age presented as mean +/- SD. LN: lymph nodes.

|  |  | Groups |  |  |  |  |
| --- | --- | --- | --- | --- | --- | --- |
| Parameters |  | Control | PCT | PCU | SeroTL | SeroCATT |
|  |  | n/total (%) or mean (SD) | n/total (%) or mean (SD) | n/total (%) or mean (SD) | n/total (%) or mean (SD) | n/total (%) or mean (SD) |
| **Epidemiological** | Age* | 32.7 (11.3) | 32.5 (16.8) | 46.6 (22.2) | 42.1 (18.1) | 40.2 (10.8) |
|  | Sex (Male) | 13/29 (45%) | 4/6 (67%) | 5/7 (71%) | 8/19 (42%) | 6/13 (46%) |
|  | Occupational risk | 23/28 (82%) | 2/6 (33%) | 3/6 (50%) | 11/19 (58%) | 11/13 (85%) |
|  | HAT case(s) in the family | 2/27 (7%) | 3/6 (50%) | 1/7 (14%) | 7/19 (37%) | 2/13 (15%) |
| **Clinical** | Headache | 15/27 (55%) | 2/6 (33%) | 2/7 (29%) | 8/19 (42%) | 4/13 (31%) |
|  | Asthenia | 12/27 (44%) | 2/6 (33%) | 2/7 (29%) | 7/19 (37%) | 5/13 (38%) |
|  | Fever | 10/27 (37%) | 2/6 (33%) | 1/7 (14%) | 5/19 (26%) | 1/12 (8%) |
|  | Any skin symptoms | 7/28 (25%) | 2/6 (33%) | 3/7 (43%) | 9/19 (47%) | 3/12 (25%) |
|  | Pruritus | 5/28 (18%) | 1/6 (17%) | 2/7 (29%) | 8/19 (42%) | 2/12 (12%) |
|  | Dermatitis | 3/28 (11%) | 2/6 (33%) | 2/7 (29%) | 3/19 (16%) | 2/12 (12%) |
|  | Swollen LN | 2/27 (7%) | 1/6 (17%) | 2/7 (29%) | 5/19 (26%) | 3/13 (23%) |
|  | Eating disorders | 2/27 (7%) | 0/6 (0%) | 0/7 (0%) | 5/19 (26%) | 0/13 (0%) |
|  | Weight loss | 2/27 (7%) | 1/6 (17%) | 2/7 (29%) | 5/19 (26%) | 2/13 (15%) |
|  | Circadian rhythm disruptions | 2/27 (7%) | 1/6 (17%) | 1/7 (14%) | 3/19 (16%) | 2/13 (15%) |
|  | Behaviour changes | 2/27 (7%) | 1/6 (17%) | 2/6 (33%) | 1/19 (5%) | 0/13 (0%) |
|  | Sexual dysfunctions | 3/27 (4%) | 0/5 (0%) | 0/7 (0%) | 3/19 (16%) | 2/13 (15%) |

One single individual was confirmed as a stage 2 case. This man in his 30’s was well-known by the medical team due to an atypical medical history: he was already diagnosed as a stage 2 case 20 years before, but at that time, his family refused the treatment. At enrolment, he was presenting swollen lymph nodes and heavy neurological symptoms, generalized pruritus and dermatitis, and he was tested CATTwb+, CATTp 1/32, RDT SD Bioline HAT+, mAECT-but CSF+ (WBC at 180), TL+, blood PCR+ but skin PCR- and IHC- (subject #1015CI in Sup. Table 1). He received a standard NECT treatment and was followed up twice. Most clinical signs resolved between the treatment and the first follow-up visit, together with the negativation of the parasitological tests. However, some neurological sequelae were still observed in 2022. His CATTp (1/8) and TL remained positive over the study and all other tests were negative excepted the PCR on skin at follow-up 1.

Two subjects died during the period of the study, yet with no additional information regarding the cause of the deaths. The first subject was a man who was only positive in TL at enrollment and follow-up 1. The second one was a woman with pruritus and dermatitis, who was tested CATTwb+, CATTp-, mAECT-, TL+, skin PCR+ and IHC+ at her last follow-up visit.

### Serological, molecular and histological test results

As compared to the control group at enrollment (0%, 0/29), 77% (10/13), 61% (8/13) and 61% (8/13) individuals in the SeroCATT group were tested positive in CATTwb, CATTp and RDT, respectively (Table 4). Positivity in CATT and RDT was also observed in the other groups, especially in treated and untreated previous cases (PCT and PCU), but this was significantly different from the control group only for PCT in CATTwb (3/6, 50%). In contrast, the positivity rates in TL for all VATs among the treated and untreated previous cases, as well as in seroTL individuals, was significantly greater than in the control group (0/29, 0%) (Table 4). No significant difference was observed between groups neither in PCR on blood nor in PCR on skin. However, 3 subjects in the SeroTL group and 1 in the SeroCATT group were found positive in PCR on skin (Table 4). The control subject tested positive in both PCR on skin and IHC was seronegative in all tests but was presenting a generalized pruritus, asthenia and headache for 2 two months (Sup Table 1). Importantly, immuno-histological analyses of skin samples revealed that parasites can be detected in all groups, and with significantly higher proportions of subjects of the PCT group (3/6, 50%) and the SeroCATT group (4/10, 40%), as compared to seronegative controls (1/29, 3%) (Table 4). In total, the presence of extravascular dermal trypanosomes was assessed by IHC (fixed trypanosome cells) and/or PCR (trypanosome DNA) on skin in about 1/3 of the cohort subjects (14/45, 31%) (Sup Table 1).

**Table 4.**
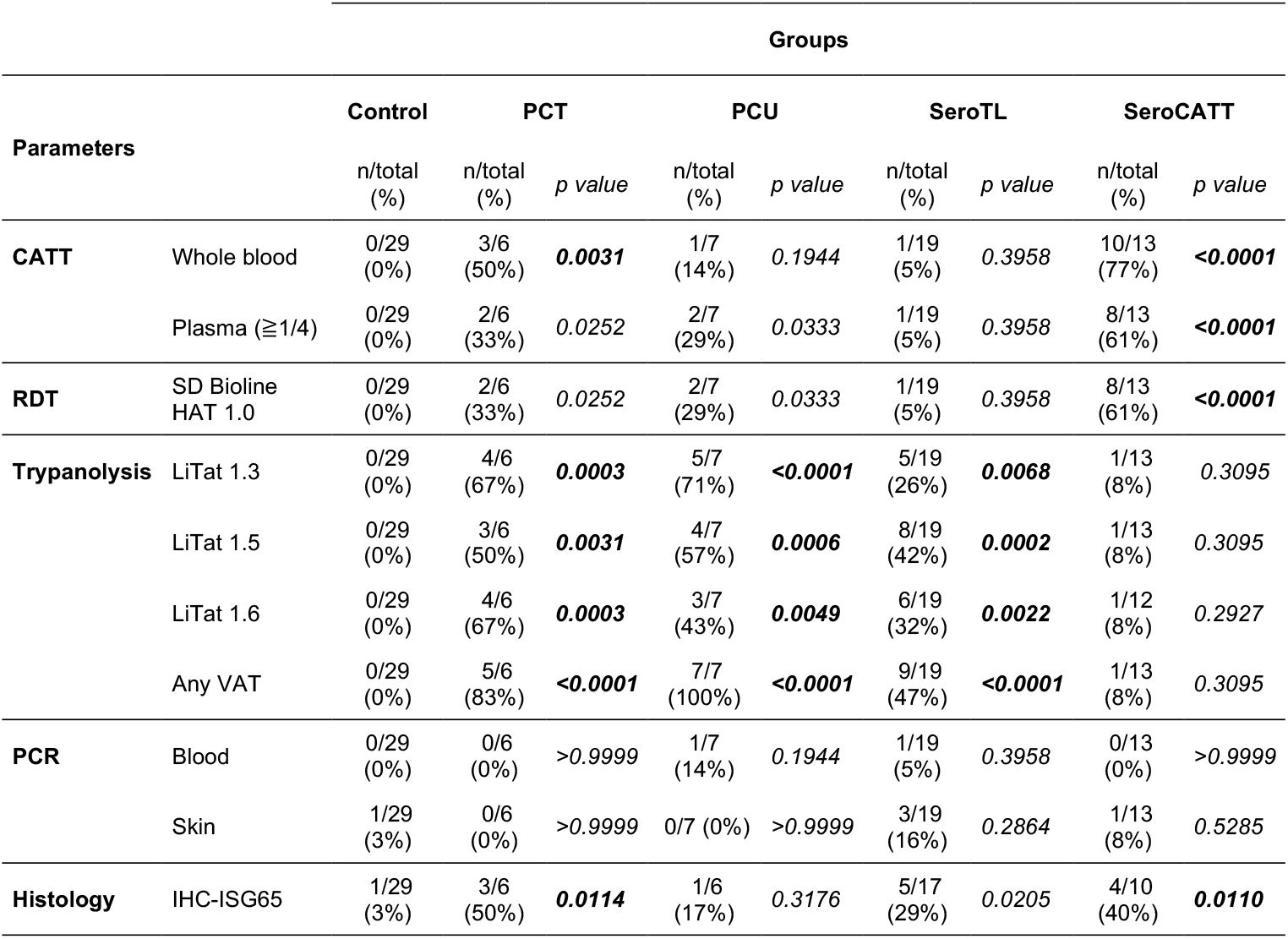
Serological, molecular and immuno-histological analysis results from blood and skin samples at enrollment. For each group (defined in Table 2) and each parameter, numbers represent the number of positive outcomes for a given test out of the total number of available results for this test. p values were obtained by comparing one by one the parameters of subjects in the seropositive Vs. seronegative groups using two-sided Fisher’s exact tests at 5% confidence. CATT: card agglutination test for trypanosomiasis; RDT: Rapid Diagnostic Test for HAT; PCR: polymerase chain reaction against TBR; IHC: immuno-histochemistry against the invariant surface glycoprotein 65 (ISG65).

Follow-up results presented in Table 5 show some positive results in all tests in all groups with variations over time, yet without clear trend, especially because of the limited number of subjects. The most striking observations over time were the persistence of positivity in TL in all previous untreated cases (PCU) and the occurrence of some positive results in PCR on skin and/or IHC in all groups.

**Table 5.** Serological, molecular and immuno-histological analysis results from blood and skin samples at follow-up visits. For each group (defined in Table 2) and each parameter, numbers represent the number of positive outcomes for a given test out of the total number of available results for this test. CATT: card agglutination test for trypanosomiasis; RDT: Rapid Diagnostic Test for HAT; PCR: polymerase chain reaction against TBR; IHC: immuno-histochemistry against the invariant surface glycoprotein 65 (ISG65).

| Test results | Groups (n/total) |  |  |  |  |  |  |  |  |  |  |  |  |  |  |  |
| --- | --- | --- | --- | --- | --- | --- | --- | --- | --- | --- | --- | --- | --- | --- | --- | --- |
|  | PCT |  |  |  | PCU |  |  |  | SeroTL |  |  |  | SeroCATT |  |  |  |
|  | 2019 | 2021 | 2022 | Total | 2019 | 2021 | 2022 | Total | 2019 | 2021 | 2022 | Total | 2019 | 2021 | 2022 | Total |
| **CATTwb +** | 2/2 | 2/2 | 0/2 | 4/6 (66%) | 1/5 | 2/5 | 0/4 | 3/14 (21%) | 1/9 | 1/9 | 1/5 | 3/23 (13%) | 7/9 | 7/9 | 4/9 | 18/27 (67%) |
| **At least one VAT+ in TL** | 1/2 | 2/2 | 2/2 | 5/6 (83%) | 5/5 | 5/5 | 4/4 | 14/14 (100%) | 6/9 | 7/9 | 1/5 | 14/23 (61%) | 1/9 | 1/9 | 0/9 | 2/27 (7%) |
| **At least one PCR + on blood** | 0/2 | 0/2 | 1/2 | 1/6 (17%) | 1/5 | 0/5 | 0/4 | 1/14 (7%) | 0/9 | 0/8 | 1/5 | 1/22 (4%) | 0/9 | 0/9 | 1/9 | 1/27 (4%) |
| **At least one PCR + on skin** | 0/2 | 1/2 | 0/2 | 1/6 (17%) | 0/5 | 3/5 | 0/4 | 3/14 (21%) | 2/9 | 3/8 | 1/5 | 6/22 (27%) | 1/9 | 3/9 | 0/9 | 4/27 (15%) |
| **At least one IHC +** | 1/2 | 1/1 | 1/2 | 3/5 (60%) | 1/4 | 4/5 | 0/4 | 5/13 (38%) | 3/7 | 6/8 | 2/5 | 11/20 (55%) | 2/6 | 6/8 | 1/9 | 9/23 (39%) |

**Table 6.** Concordance between serological test and molecular test results. The concordance between two tests represents the proportion of common negative and positive results out of the total number of reactions performed in both tests on all samples from all subjects at all timepoints. TL: immune trypanolysis test on any variable antigen type; CATTp: card agglutination test for trypanosomiasis on plasma; IHC: immuno-histochemistry against invariant surface glycoprotein 65 (ISG65); Mol Biol on skin: molecular biology test (PCR-TBRN3 and SHERLOCK) on skin samples.

| No. double positive results (%) | At least one VAT + in TL | CATTwb + | CATTp >1/4 | At least one PCR + on blood | At least one PCR + on skin | At least one IHC + |
| --- | --- | --- | --- | --- | --- | --- |
| **At least one VAT + in TL**<br>(n=43) | x | 8/43 (19%) | 10/43 (23%) | 3/42 (7%) | 6/42 (14%) | 16/37 (43%) |
| **CATTwb +** (n=32) | 8/32 (25%) | x | **23/32 (72%)** | 1/32 (3%) | 10/32 (31%) | 12/28 (43%) |
| **CATTp ≥1/4** (n=30) | 10/30 (33%) | **23/30 (77%)** | x | 2/30 (7%) | 8/30 (27%) | 9/26 (35%) |
| **At least one PCR + on blood** (n=5) | 3/5 (60%) | 1/5 (20%) | 2/5 (40%) | x | 0/5 (0%) | 2/5 (40%) |
| **At least one PCR + on skin** (n=16) | 6/16 (37%) | 10/16 (62%) | 8/16 (50%) | 0/16 (0%) | x | 10/16 (62%) |
| **At least one IHC +** (n=35) | 16/35 (46%) | 12/35 (34%) | 9/35 (26%) | 2/35 (6%) | 10/35 (29%) | x |

## Discussion

To refine our observations of extravascular dermal trypanosome carriage [10, 11], we investigated whether *T. b. gambiense* parasites might also be found in the skin of seropositive suspects in hypo-endemic foci in Côte d’Ivoire.

Here, we observed a non-negligeable proportion (about 1/3) of unconfirmed seropositive subjects with extravascular trypanosomes detected in the skin by molecular biology (trypanosome DNA) and/or immuno-histochemistry (trypanosome cells) approaches, demonstrating that this phenomenon is not restricted to the active transmission foci in Guinea. However, the frequency of dermal trypanosome carriage observed at enrolment in seropositive suspects was apparently lower than in Guinea, and more importantly, not clearly associated neither with dermatological signs nor with serological positivity in any test. As compared to previous case/control studies [10, 11, 20], the unusual and heterogeneous composition of the cohort followed-up in the present study likely explains these intriguing observations. Indeed, the cohort includes 45 subjects who were either previously treated HAT cases, ancient non-treated HAT cases (treatment refusals) or long-lasting seropositive individuals, with the common point of being detected with a variable seropositivity over several years of follow-up. Yet, in the present study, no correlation was observed between CATT or TL and positivity in dermal parasite detection by molecular methods. Could this result from a particular immune response in the dermal anatomical niche of these subjects with immunogenetic specificities?

Among PCU, only 17% and 0% were found positive in IHC and PCR on skin, respectively. These results could be interpretated as self-cures, yet, as discussed previously [11], the proportion of skin samples found positive for trypanosomes (DNA or fixed cells) could rather likely be under-estimated due to the superficial nature of snip biopsies that necessarily contain less dermal tissue than punch biopsies, and therefore potentially less dermal trypanosomes that were shown to be enriched in the basal dermis [8]. As there exist only few reports and no gold-standard approach for detection of trypanosomes in the human skin [10, 11, 21], we implemented complementary molecular and immuno-histological methods in parallel. As discussed previously, these methods have their own specific strengths and weaknesses [10]. Considering that *T*.*b. brucei* are theoretically non-infectious to humans and are killed within a couple of hours by human serum *in vitro* [22], the dermal parasites detected here are likely to be *T*.*b. gambiense* parasites, although the well-known low sensitivity of the TgsGP-PCR reference assay targeting a single copy gene did not allow us to confirm their identity in this study [23, 24]. This questions the exact identity of the skin-dwelling trypanosomes that are found in all groups studied here. They are detected in subjects with negative trypanolysis tests (seroCATT), yet their presence is apparently also linked to positivity in CATT and/or RDT in the other groups, which also emphasizes the need to better understand the underlying immune mechanisms.

As recently observed in Cameroon [25], one possible explanation for the persistence of disease foci in certain regions is the presence of animal reservoirs carrying trypanosomes in both blood and skin [26]. Another possibility is that traditionally used diagnostic approaches based on blood examination do not detect some *T*.*b. gambiense* infections among seropositive cases [26]. Mathematical modelling predicted that, in the absence of any animal reservoirs, these unconfirmed seropositive individuals could contribute to disease transmission by maintaining an overlooked reservoir of skin-dwelling parasites, which would slow down progress towards elimination [27]. The phenomenon of human trypanotolerance, that was first described in Côte d’Ivoire in an initial cohort of about 50 seropositive subjects [7], could be reinforced here, by the detection of skin-dwelling trypanosomes in some of these individuals with no parasite detectable in the blood. Nevertheless, considering the limited number of seropositive individuals, the duration of vector control activities in the studied regions, and in absence of confirmed cases, their potential role in contributing to inter-human transmission is likely limited in comparison to the situation observed in Guinea [28].

In total, these results raise questions about the strategies used to diagnose this disease as the detection of skin-dwelling trypanosomes would allow more carriers to be treated. The current WHO recommendation, based on risk-benefit analyses, is to not treat unconfirmed seropositive individuals without knowing if they have an active infection [29]. With the promise of Acoziborole, a new cheaper and less toxic drug that only requires a single oral administration, the policy of treating unconfirmed seropositive individuals bearing dermal trypanosomes could possibly be reconsidered [30]. This would certainly benefit the epidemiological transition to zero incidence and the sustained elimination of transmission.

## Data Availability

All data are provided

## Acknowledgements

We thank M. Carrington (University of Cambridge, UK) and J. Bangs (University at Buffalo, USA) for providing antibodies. We warmly thank the team of the Programme National d’Elimination de la Trypanosomiase Humaine Africaine of Côte d’Ivoire, the Institut Pierre Richet of Bouaké and the University Jean Lorougnon Guédé of Daloa, as well as all our collaborators of the Sinfra and Bonon Health Departments.

## Author contributions

- Investigation: Martial Kassi N’Djetchi, Mélika Barkissa Traoré, Innocent Abé, Bamoro Coulibaly, Valentin Nanan, Thomas Konan, Louis N’Dri, Ibrahim Sadissou, Jean-Mathieu Bart, Bruno Bucheton, Magali Tichit, Salimatou Boiro, Aïssata Camara, Christelle Travaillé, Aline Crouzols, Nathalie Petiot, Adeline Ségard, Mariame Camara, Mathurin Koffi, Vincent Jamonneau and Brice Rotureau
- Supervision and logistics: David Hardy, Lingue Kouakou, Mamadou Camara, Dramane Kaba, Mathurin Koffi, Vincent Jamonneau and Brice Rotureau
- Data curation and formal analysis: Vincent Jamonneau and Brice Rotureau
- Original Draft Preparation: Brice Rotureau
- Review & Editing: Martial Kassi N’Djetchi, Bruno Bucheton, Vincent Jamonneau and Brice Rotureau

---

**Supplementary Table 1. Raw data used in the study.**

Empty cells correspond to absent data (information / sample not collected or test not done), 0 for negative occurrence / result and 1 for positive occurrence / result. M: male; F: female; ID: anonymized subject identifier; LN: lymph nodes; CATTwb / CATTp: card agglutination test for trypanosomiasis on whole blood / plasma; RDT: Rapid Diagnostic Test for HAT; mAECTbc: mini anion-exchange column technique on buffy coat; WBC: white blood cells; TL: trypanolysis immune test; PCR: polymerase chain reaction against TBR; IHC: immuno-histochemistry against the invariant surface glycoprotein 65 (ISG65). Enrolment in 2019, first follow-up visit in 2021 and second in 2022.

